# The local tumor microbiome is associated with survival in late-stage colorectal cancer patients

**DOI:** 10.1101/2022.09.16.22279353

**Authors:** Justine W. Debelius, Lars Engstrand, Andreas Matussek, Nele Brusselaers, James T. Morton, Margaretha Stenmarker, Renate S. Olsen

## Abstract

**Objective:** Colorectal cancer survival has been linked to the microbiome. Single organism analyses suggest *Fusobacterium nucleatum* as a marker of poor prognosis. However, *in situ* imaging of tumors demonstrate a polymicrobial tumor-associated community. To understand the role of these polymicrobial communities in survival, we performed an untargeted study of the microbiome in late-stage colorectal cancer patients.

**Design:** We conducted a nested case-control study in late-stage cancer patients undergoing resection for primary adenocarcinoma. The microbiome of paired colorectal tumor and adjacent tissue samples was profiled using 16S rRNA sequencing; we used compositionally aware ordination and differential ranking to profile the microbial community.

**Results:** We found a consistent difference in the microbiome between paired tumor and adjacent tissue, despite strong individual microbial identities. Tumors had higher relative abundance of genus *Fusobacteria* and *Campylobacter* at the expense of members of families Lachnospriaceae and Rumminococeae. Furthermore, a larger difference between normal and tumor tissue was associated with prognosis: patients with shorter survival had a larger difference between normal and tumor tissue. We found the difference was specifically related to taxa previously associated with cancer. Within the tumor tissue, we identified a 39 member community statistic associated with survival; for every log2 fold increase in this value, an individual’s odds of survival increased by 20% (OR survival 1.20; 95% CI 1.04, 1.33).

**Conclusion:** Our results suggest that a polymicrobial tumor-specific microbiome is associated with survival in late-stage colorectal cancer patients.

## INTRODUCTION

Globally, colorectal cancer (CRC) is the second most common cause of cancer-related death and CRC-related mortality has been increasing since 2000 (1,2). One potentially modifiable area of interest in CRC survival is the gut microbiome. In a healthy gut, the intestinal microbiome contributes to homeostasis through epithelial cell renewal, maintaining gut barrier integrity, and immune modulation (3,4). However, CRC patients have demonstrated a consistently altered gut microbiome compared to healthy controls, including a higher relative abundance of organisms more commonly found in the oral cavity (5,6). Meta-analyses using targeted analyses show high levels of *Fusobacterium nucleatum* (*F. nucleatum*) in tumor tissue are detrimental to survival (7,8).

Fewer studies have explored the relationship between the gut microbiome and CRC prognosis using untargeted sequencing. Untargeted techniques can better characterize the bacterial community, and the ways in which potentially pathogenic organisms might interact with a host’s unique, stable, microbiome (9–11). *In situ* microscopy shows that tumor tissue is colonized by apolymicrobial biofilm including Fusobacteria, Proteobacteria, Bacteroidetes, and Lachnospriaceae; monoculture biofilms have not been observed (12). Biofilms are also frequently localized to tumors, and paired normal tissue samples are rarely colonized, suggesting a localized effect and potential difference between tumor and adjacent tissue (12).

Previous untargeted studies of the gut microbiome and colorectal cancer survival have either focused exclusively on the tumor tissue (13) or have treated the tumor and adjacent tissue as identical (14). Paired biopsy studies provide clues about whether local and global regulation of the microbiome drives tumorigenesis, although many paired studies have failed to account for survival (12,15–19), and in some cases, struggled to characterize the microbiome due to technical (19) or analytical (13–17) issues.

To address the gaps in knowledge, we followed 101 late-stage CRC patients recruited from a hospital in southern Sweden who underwent surgical resections of primary adenocarcinoma between 1997-2017. Patients were categorized into short- or long-term survivors based on their relapse free survival (less than two years or five or more years, respectively). We examined the relationship between the microbiome of colorectal tumors and adjacent normal tissue and survival, accounting for clinical covariates.

## METHODS

### Study Population

Patients were recruited from all consecutive CRC patients (n=540) who underwent surgical resection for primary colorectal adenocarcinoma at the Department of Surgery, Ryhov County Hospital, Region of Jönköping County, Jönköping, Sweden between 1997 and 2017. Patients with tumor-node-metastasis (TNM) stage III and IV cancer who had matched biopsies from normal and tumor tissue (n=116) were selected. Patient characteristics, including demographic, surgical, pathological information, and outcome were determined from a review of medical records.

The final study cohort included patients with paired high quality microbiome samples (n=101). Fifteen individuals were excluded due to insufficient sequencing depth in the tumor (n=8) or normal (n=7) tissue sample. There was no difference in the survival status in the samples with insufficient sequencing depth. Our final cohort included matched tumor- and normal tissue samples (≥10 cm apart from tumor tissue) from 51 long- (≥5 year survival) and 50 short-term (≤2 year survival) survivors.

The study was approved by the Regional Ethical Review Board in Linköping, Linköping, Sweden (98113, 2013/271-31); a written informed consent was obtained from each patient.

There was no patient or public involvement in this retrospective study.

### Statistical Analysis of patient characteristics

A multivariable logistic regression was used to assess the predictive impact of the following patient, cancer and treatment related characteristics: age (categorized as <60, 60-69, 70-74, ≥75 years), sex (female or male), American Society of Anesthesiologists physical status (ASA) score (I-healthy, II-mild, III-IV-severe; patients with V-VI were not eligible for surgery); localization of tumor (colon right, colon left, rectum), TNM stage (III or IV), grade of differentiation (from low differentiation to high differentiation, with the latter more closely resembling non-cancer histology); radical surgery (yes or no); period of surgery (1997-2005; 2006-2010; 2011-2017). All results are expressed as Odds Ratios (ORs) and 95% Confidence Intervals (CIs) and the calculations were conducted with Stata MP14 (Stata Corp, Texas).

### Microbiome Sequencing

Paired tumor and normal tissue samples were collected during surgery. Tissue samples were frozen directly and stored at -80 °C until use. Samples were processed as previously described (20). Briefly, DNA was extracted from tissue samples using physical and chemical lysis for extraction. The 16S rRNA amplicon library was amplified with 341F/805R primers (CCTACGGGNGGCWGCAG, GGACTACHVGGGTATCTAAT) using a program with 20 cycles (21). The samples were sequenced with a 2×300 approach using an Illumina MiSeq (San Diego, CA, USA).

The demultiplexed reads were denoised using the DADA2 algorithm (v1.13.1) in R (22). After reads were demultiplexed and primers were trimmed, forward reads were trimmed to 265 nucleotides (nt) and reverse reads were trimmed to 225nt; the error rate model was trained on 15% of the reads. Reads were joined with at least 30nt overlap, and anything shorter than 380nt after joining was discarded. Taxonomic assignment was performed using the naïve Bayesian classifier implemented in DADA2 against the Silva 128 database (23,24). The amplicon sequence variant (ASV) table from DADA2, taxonomy, and representative sequences were imported into QIIME 2 (v. 2020.11) for further processing (25). A phylogenetic tree was built using fragment insertion using the SEPP algorithm into the Silva 128 backbone with q2-fragment-insertion (24–26). The table and sequences were filtered to exclude any ASV without phylum level annotation or which could not be inserted into the phylogenetic tree.

### Microbiome Community Characterization

#### Between sample (beta) diversity

For paired sample analysis, we calculated unweighted UniFrac (27), weighted UniFrac (28), and binary Jaccard (29) distances and Bray-Curtis dissimilarity (30) on a feature table rarified to 2500 sequences/sample (31). Aitchison distance was calculated on unrarefied data with a pseudo-count of 1 (32,33). Beta diversity metrics were calculated using the q2-diversity plugin in QIIME 2 (25).

#### Compositional Tensor Factorization ordination

To account for subject-specific effects on ordinations, we used compositional tensor fraction (CTF) for paired samples using the Gemelli qiime2 plugin (0.7.0) (34). Features were filtered to exclude those present in fewer than 20 samples or less than 100 total counts. The distance in CTF subject space was calculated as the Euclidean distance between subject coordinates. The difference in intra-individual CTF space between normal and tumor tissue (ΔPC) were compared using the subject-state coordinates and feature-state coordinates, respectively.

#### Robust Principal Components Analysis

For each tissue type, we examined beta diversity using a Robust Principal Components Analysis (rPCA) using the DEICODE algorithm (v. 0.2.4) (35). For a given sample set, we filtered filtering features present in less than 10% of samples (n=10) or with fewer than 10 total counts. The auto-rPCA function was used to select the appropriate number of principle components (PCs) for the data. The PCs were divided into quartiles and dichotomized along the median value.

For tissue types where there was a significant association between a component and survival, we selected features which might be associated with outcome. Communality was calculated as the square root of the sum of squares across all PC. Features with a communality value of at least 0.01 were selected as candidates for the additive log ratio (ALR) calculation (n=130). A pseudocount of 1 was added before the ALR calculation. The ALR was calculated as the log2 ratio of features more extreme that the fourth quartile of samples over feature more extreme than the first quartile. Continuous ALR values or ALR divided into tertials were used for regression.

#### Differential Ranking

We performed hypothesis generating differential abundance testing between tumor and normal tissue using a modified differential ranking (DR) technique (36,37). We first filtered the table to remove any feature with a relative abundance of less than 1/1000 in fewer than 10% of samples, leaving 243 features for testing. We then used a modified Bayesian method for DR testing. ASV counts were modeled through a negative binomial process. We started with naive priors of a between 0-and 5-fold change in a ASV and fit the model using 4000 iterations. The data was fit to a linear mixed effects model using subject as a random intercept, modeling either for tissue or for the intersection between tissue and survival. Modeling was done with pystan (v 3.4.0) within the QIIME 2 2021.11 conda environment (38,39).

We used the ranks to identify “extreme” features. Starting from the feature with the strongest signal associated with each possible value for a variable (e.g. normal vs tumor, short vs long survival), we added features until every sample contained at least one of the extreme features. A pooled ALR was calculated as the sum of all normal-tissue associated features over the tumor-associated features.

### Statistical Analysis

Paired distances were extracted as the distance between an individual’s tumor and adjacent normal tissue. Interindividual distance was compared to the interindividual distance to samples of the same tissue type, anatomical location, and survival group with a permutative two sample t-test with 999 permutations.

Associations with per-subject CTF coordinates were checked by calculating the Euclidean distance between samples and applying a permanova test with 999 permutations in scikit-bio (v. 0.5.6) (40). The change between tissue types in CTF coordinate space were modeled with a paired sample t-test was used to determine if there was a global difference between tumor and normal tissue along either PC; the effect of change on survival was compared using a permutative Welch’s t-test looking at the difference between groups with 999 permutations. ALR interactions were evaluated using a linear mixed effects model with individual as the grouping factor.

Survival was modeled using logistic regression. Models were fit using a crude (unadjusted) model and a model adjusted for age, sex, ASA score, tumor location, surgery period, TNM stage, radical surgery, and differentiation grade.

For all analyses, a p-value of 0.05 was considered significant.

Modeling was performed using statsmodels (v 0.11.1), scipy (v 1.4.1), and numpy (v 1.18.5) in python (v. 3.6) (41–43). Figures were plotted using with matplotlib (3.2.2) and seaborn (0.10.1) The dendrogram was plotted using Empress (q2-empress v.0.0.1-dev, commit b705358) (44); three dimensional ordinations were rendered using Emperor (v. 1.0.3) (45). Taxonomic colors come from the microshades colorblind friendly palette (46). Figures were assembled in Adobe Illustrator 2021 (Adobe Inc, San Jose, CA, USA).

## RESULTS

In our nested case-control study of late-stage colorectal cancer patients, the 51 long-term survivors were more likely to be younger, male, and healthier compared to the 50 short-term survivors (Table S1). The short-term survivors presented with metastatic tumors and lower differentiation than long-term survivors, and fewer received radical surgery. We found that age, TNM stage and tumor differentiation were strong predictors of long-term survival (Table 1). Individuals aged between 70-74 years were 14 times more likely to be short-term survivors (OR 14.24; 95% CI 1.21, 167.40) than those younger than 60. TNM-stage IV was associated with an almost 50 times higher risk of being a short-term survivor (OR 49.32; 95% CI 5.86, 415.12) compared to TNM stage III (Table 1).

**Table 1.**
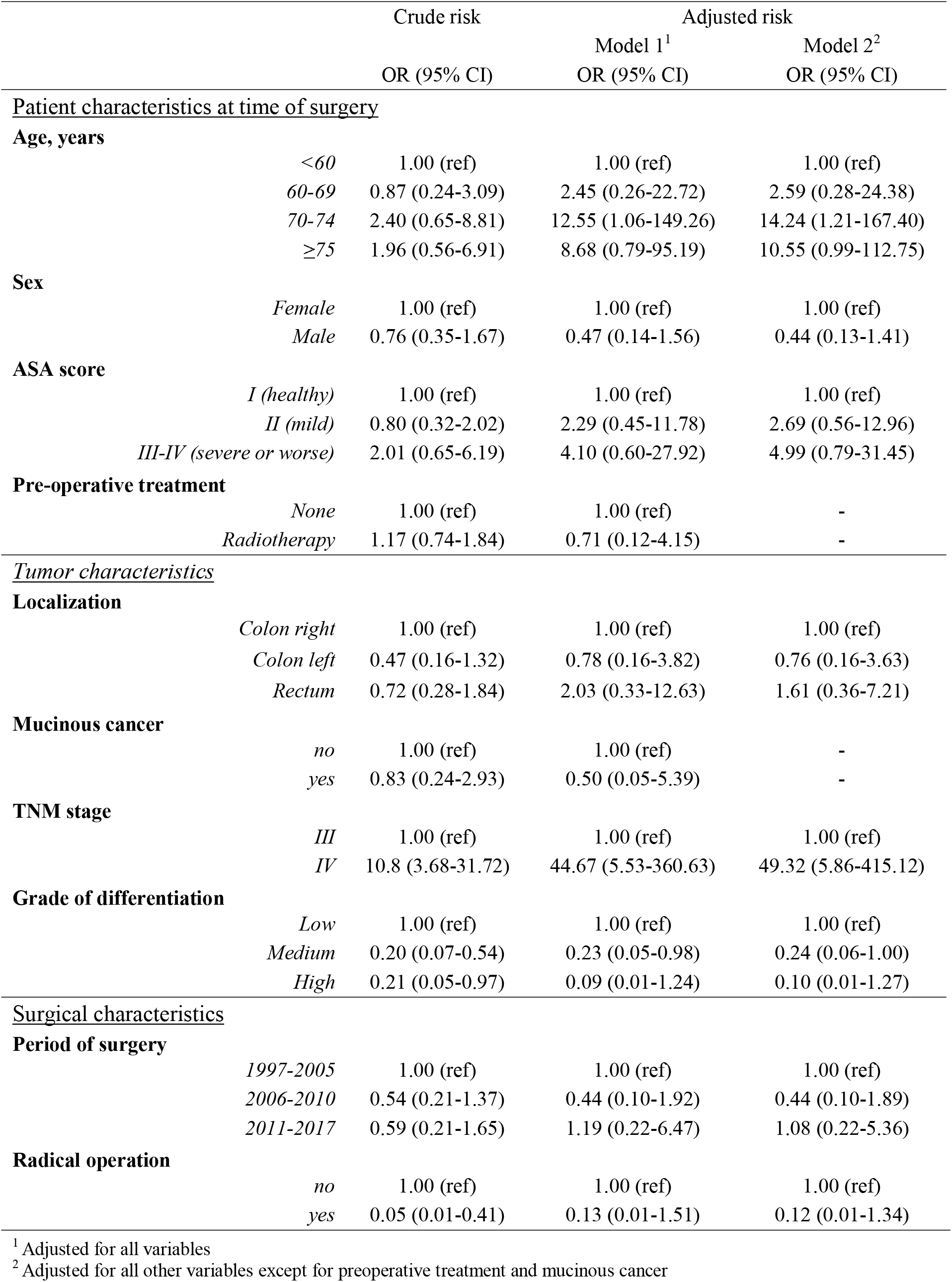
Risk factors for short-term survival.

Following sequencing, quality filtering, and denoising to ASVs, we retained 202 paired tumor and adjacent normal tissue samples. The broad pattern in the overserved microbiome reflect those seen in previous studies of Swedish adults (Figure S1) (47). We found the patient was the strongest predictor of microbiome composition, and that an individual’s paired samples were more similar to each other than tissue samples from patients with similar characteristics (Figure S2), reflecting what appears to be a common pattern in CRC patients and beyond (10,18,48).

### The microbiome of tumor and normal tissue differ

To address individual microbial identities, we applied a subject-aware compositional tensor factorization (CTF) technique (34). The approach integrates feature-based information from each sample to build both a subject-specific profile, and to describe changes within subjects and features across a gradient. We did not find a statistically significant association between a sample’s position in CTF space and survival (unadjusted permanova R^2^=0.012; p=0.296, 999 permutation, Figure S3, Table S2). However, we found differences between normal and tumor tissue, with a consistent shift between paired samples in CTF space, primarily along principal component (PC) 2 and PC 3 (Figure 1A-D).

**Figure 1.**
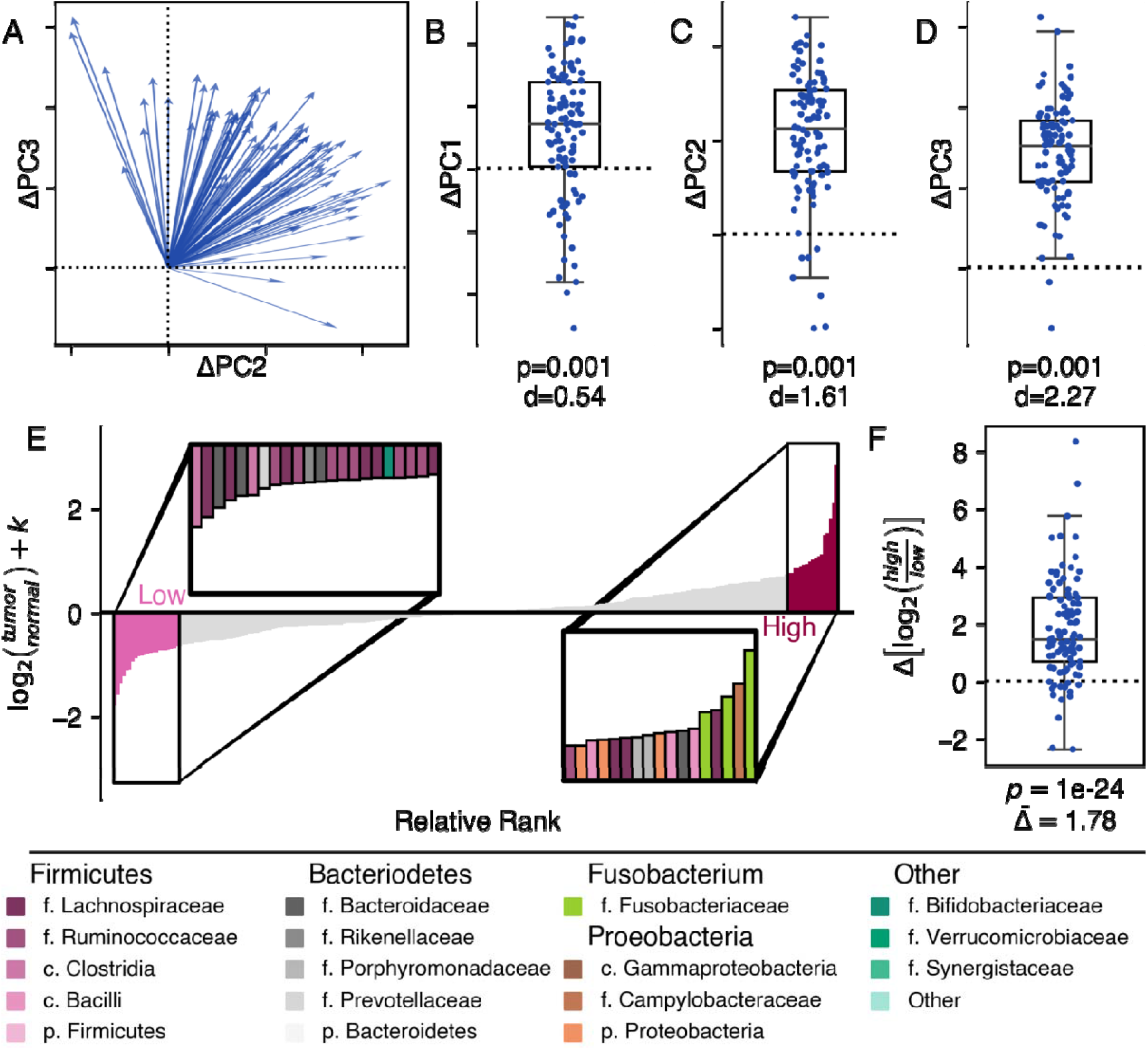
There is a difference in the microbiome between tumor and normal tissues. We found a global pattern separating tumor and normal tissue can be seen in CTF ordination space. (A) Plotting the change between normal tissue and tumor tissue in PC 2 and PC 3 as a vector with normal tissue as the center demonstrates a clear directional pattern. The difference between normal and tumor tissue can also be seen along individual components: (B) PC 1, (C) PC 2, and (D) PC 3. Ticks and dashed zero-lines along PC 2 (B) and PC 3 (D) match the two-dimensional axes in (A). All boxplots are shown with a Cohen’s d effect size statistic for a one-sided t-test and p-values from a permutative one sample t-test, 999 permutations. (E) Differential ranking of 300 abundant features identified normal tissue-associated features (light pink) and tumor-tissue associated features (dark pink). The inset shows selected features in each group, colored by family, colors are defined in the legend. (F) The change in the ALR between normal and tumor tissue. Coefficient from linear mixed effect model comparing the change based on tissue type; p < 1×10^−12^.

Given evidence of consistent, community-level changes in the microbiome between the tissue types, we looked for features, which might be driving these differences. We used an individual-aware differential ranking technique, which first ranked the features with the greatest differences associated with tissue type, and then we selected a subset of these features to build an additive log ratio (ALR), a summary of taxa which likely describe the difference (Figure 1E, Table S3, Files S1). We found tumor tissue was associated with a higher relative abundance of *Fusobacteria, Porphyromonas, Granulicatella*, and *Campylobacter* at the expense of members of genus *Blautia*, and *Ruminococcus*. Tumor tissue had a 1.78 (95% CI 1.50, 2.18, p < 1×10^−12^) log2-fold increase in the features selected by DR compared to normal tissue, suggesting a tissue-specific signature (Figure 1F).

### Differences between normal- and tumor-associated microbiome are associated with survival

Since we saw consistent differences between tumor and normal tissue, we wondered if there might be a relationship between the magnitude of the difference and survival. We found that tumor and normal tissue were more similar in long-term survivors than short-term, a difference primarily driven by changes in abundance (Table S4). Additionally, long-term survivors showed a larger change along PC 2 compared to short-term survivors (Cohen’s d 0.40, p=0.016, 999 permutations; Figure 2). This suggested enough of a community-level change in the microbiome to motivate looking for features which might explain the differences.

**Figure 2.**
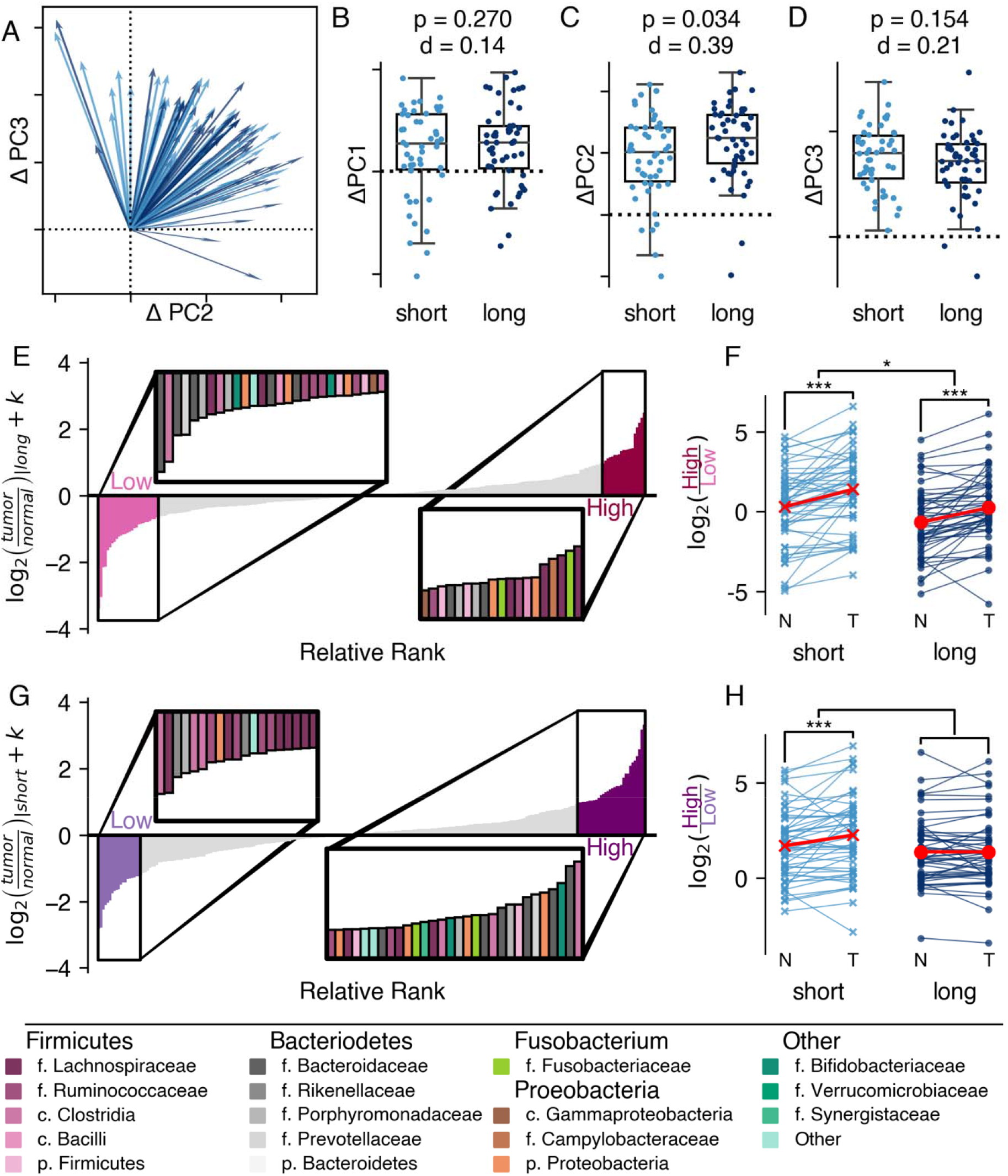
The magnitude of the difference between tumor and normal tissue is associated with survival. There is a difference in the magnitude of change between long-term and short-term survivors. (A) In two dimensions, the change along PC2 and PC3 is visualized as a vector from normal tissue to tumor issue. Short-term survivors (<2 year) are shown in light blue. Long term survivors (≥5 years) are shown in dark blue. The corresponding relationships can be visualized along the individual components: (B) PC 1, (C) PC 2, and (D) PC 3. Ticks along PC 2 (C) and PC 3 (D) match the two-dimensional axes in (A). All boxplots are shown with a Cohen’s d effect size, and p-value from a permutative Welch’s t-test with 999 permutations, comparing the two survival groups. (E-H) A differential ranking model was fit to consider the interaction between survival and tissue. The ranks associated with (E,F) tumor tissue in long-term survivors and (G,H) tumor tissue in short survivors (interaction) are show. (E,F) The relative associated with the model, insets highlight ASVs associated with the extremes of each group. Taxonomic assignments are provided in the legend. (F,H) The additive long ratio associated with the ranks. Paired differences are connected by a line between normal (N) and tumor (T) tissue. The effect was modeled using a linear mixed effects model, treating the individual as random. *p < 0.05, **p ≤ 0.01, ***p≤0.001.

Therefore, we applied a subject-aware differential ranking technique looking at the interaction between tissue type and survival to further refine the features (Figure 2E-G). We used an interaction model to identify features that changed in tumor tissue based on survival group. Based on the tissue associated taxa associated with long-term survival, we defined an ALR where tumor tissue was associated with ASVs from genus *Fusobacterium, Campylobacter*, and *Escherichia/Shigella*. We found members of genus *Butyricicoccus, Roseburia*, and *Streptococcus* associated with both normal and tumor tissue (Table S5, File S2). There was a higher relative abundance of the tumor-associated organisms in both survival groups, and the overall relative abundance was higher in short-term survivors (Figure 2F). However, the magnitude of the change did not differ between the two groups.

In contrast, the interaction term identified a set of taxa, which were significantly different between the tissue types in short-term survivors but not among long-term survivors (Figure 2G, Table S6, File S2). Once again, we found tumor tissue in short-term survivors to be strongly associated with an ASV from *Fusobacteria* and as well as a few members of family Veillonellaceae, although again, there were not clear taxonomic patterns in other families, such as family Lachnospiraceae or Rumminococeae. These results indicate the survival-associated changes in the microbiome may be largest in tumor tissue and help to identify specific set of organisms responsible for these changes.

### The tumor microbiome is associated with survival

Based on our observation that differences in tissue types were more pronounced in short-term survivors, and since past work focused on tumor tissues, we also chose to further interrogate the tumor-specific microbiome. A rPCA approach showed separation in the microbiomes between short- and long-term survivors (Figure 3A, D) (34). After adjustment for confounders and both PCs, patients with larger values for PC 1 had 3.5 lower (OR 0.29; 95% CI 0.08, 0.97) odds of short-term survival, while those with higher values for PC 2 were five times less likely to be short-term survivors (OR 0.19; 95% CI 0.05, 0.80). Individuals in the quadrant defined by these two extremes in the data were at least 7.5 times more likely to survive than any other group in the ordination (Figure S4).

**Figure 3.**
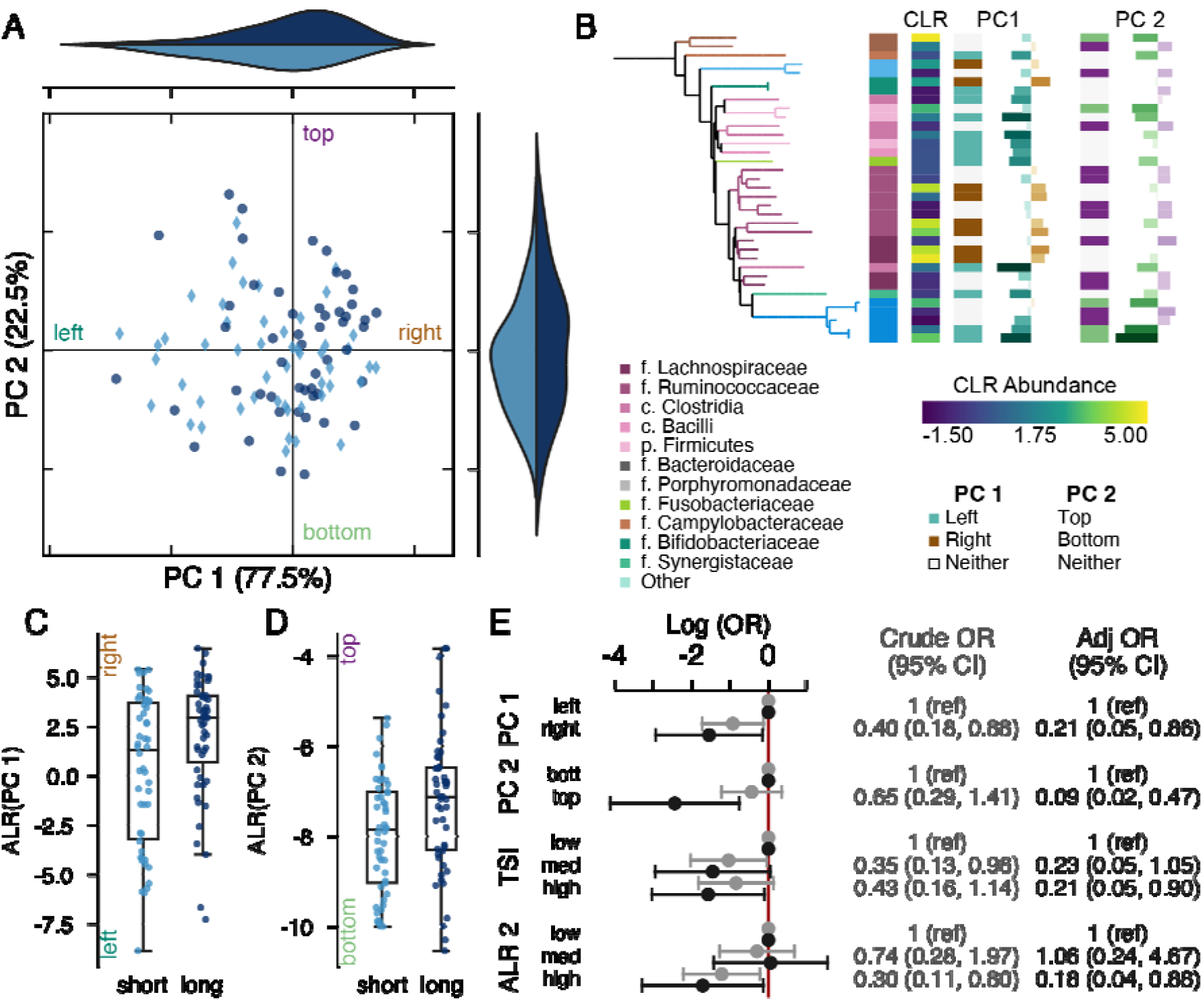
The tumor-associated microbiome is associated with survival. (A) Robust Principal Components Analysis (rPCA) ordination colored by short- (light) or long-(dark) term survival. Marginal axes show the distribution of points along each PC. The ordination is centered at the median distribution of points in each axis. (B) Phylogenetic tree showing the ASVs with PC 1. The tips and first heatmap are colored with taxonomic information. Heatmaps from left to right show the taxonomic assignment, mean central log ratio (CLR) relative abundance (viridis), the mean difference in CLR between long- and short-term survivors; whether the feature was used in the additive log ratio (ALR) calculation for PC 1 (teal - left; brown - right); the feature loading along PC 1; whether the feature was used in the PC 2 ALR calculation (green - bottom, purple top); the feature loadings along PC 2, coordinates to the top on the left; and whether the feature was included in the tumor survival index (light pink - lower value, dark pink - higher value). (C,D) Boxplot of the ALR of most extreme ranked taxa along selected based on (C) PC 1 and (D) PC 2. Axes are labeled to indicate the directionality of the log ratio calculation, label colors match panel (B). (E) Log odds of survival based on separation along the median of the PCs or grouping in the ALR shown in Light gray values are crude, dark gray are adjusted for age, sex, surgery year, tumor location, ASA score, differentiation grade, TNM stage. PC weights are adjusted for the position along the other PC; ALRs are not co-adjusted.

We found 37 features associated with separation along PC 1. To the left of PC 1, we found members of genus *Fusobacterium, Parvimonas* and *Porphyromonas*, and other common oral genera like *Gemella* and *Dialster* (Figure 3B). In contrast, higher values along PC 1 (to the right) were correlated to more common gut taxa, including members of families Lachnospiraceae and Rumminococceae. We defined the log2 fold ratio between the organisms separating PC 1 as a tumor-survival index (Table S7, File S3). For every 2-fold increase in this index in tumor tissue, the odds of survival increased by 20% (adjusted OR 0.80; 95% CI 0.67, 0.96). There were no clear patterns in the taxa separating along PC 2, beyond the association between Escherichia/Shigella and short-term survival (Table S8, File S3), although there was a significant relationship between these selected taxa and survival (OR 0.64; 95% CI 0.41, 0.98 for every log2 increase).

## DISCUSSION

Our results show a clear and consistent difference between normal and tumor tissue once we had accounted for individual microbiome effects. Across all patients, tumors carried a higher proportion of ASVs mapped to genus *Fusobacterium, Gemella, Dialster* and *Campylobacter* at the expense of genera like *Blautia* and *Allistipes*. The tumor-associated features reflect organisms found more commonly in CRC patients compared to healthy controls, whereas the organisms associated with normal tissue belong to clades commonly associated with short chain fatty acids and widely believed to be beneficial (5,6,49–51). Further, we are among the first to show that the magnitude of the difference between the two tissue types can be associated with prognosis. Our differential ranking analysis identified a set of 38 ASVs, which changed between the tumor and normal tissue in short-term survivors, but not long-term survivors. This suggests survival may be associated with localized changes in the microbiome.

We are among the first to report differences a different between tumor and normal tissue in paired samples, let alone an association between the degree of dissimilarity and survival. Drewes et al (12) demonstrated clear difference between paired tumor and normal tissue samples using microscopy, although their 16S analysis did not explicitly test paired samples. These results seemingly conflict with much of the existing literature (15–18,48). Several previous studies reported no difference in the microbiome between the two tissue types, let alone an intra-individual difference associated with survival. Like the past studies, we observed and described a strong intraindividual similarity. A personal microbial signature is a normal feature of the microbiome seen in a variety of settings including population-based studies (10), dietary interventions (52), and among CRC patients (15–18,48). However, unlike prior work, the statistical models we selected accounted for this strong intra-individual similarity. Re-analysis of prior publications using subject aware methods may identify the same patterns we found: strong individual microbial signatures with a difference between the tissue types. Our results indicate that the tumor-specific microenvironment, rather than the overall microbiome, is important for understanding CRC pathology. At a minimum, future sequencing survey studies will need to account for tissue-specific effects in their analysis, and studies treating tumor and non-tumor biopsy samples as identical may need to check for biases.

Based on the difference in the microbiome between tissue types, we specifically focused on the relationship between the tumor microbiome and survival. Two previous studies have explored the relationship between the tumor microbiome and survival using untargeted sequencing. In that study of 67 Irish patients, Flemer and colleagues defined microbiome groups using a non-compositional abundance-based clustering approach (14). They found a higher relative abundance of a cluster defined by members of genus *Bacteriodetes, Blautia, Roseburia, Rumminococus*, and an unclassified member of family Lachnospiracae was associated with shorter survival, while higher abundance of a cluster characterized by *Streptococcus, Fusobacterium*, and unclassified family Enterobacteraceae was associated with longer survival. These groupings are contradictory to the features associated with survival in our tumor tissue results. In contrast, our tumor survival index, defined by an ALR of features along PC 1, showed a decrease in the relative abundance in *Fusobacterium* among our long-term survivors, who were characterized by a higher relative abundance of *Blautia, Roseburia*, among others. It is likely this disagreement is due to differences in methods used for differential abundance (36,53). Our results are more in line with results from a Chinese cohort (13). In that study, a higher untransformed relative abundance of genus *Fusobacterium*, or higher relative abundance of reads mapped to *Bacteriodetes fragilis* were associated with an increased hazard of death, while a higher relative abundance of genus *Faecalibacterium* was protective. We find similar trends in our tumor survival index, where short-term survival was associated with ASVs mapped to genus *Fusobacterium* and a *Bacteriodetes* ASV, while longer survival was associated with *Faecalibacterium*. Our results and those of the Chinese cohort suggest that a more normal (gut-like) microbiome is associated with long-term survival, while a more disrupted (oral) microbiome led to a poor prognosis.

Our conclusions are supported by our nested case-control design, which helps establishing temporality: changes in the local tumor microbiome at the time of surgery are associated with future outcomes, increasing the probability that the observation is a real phenomenon, rather than a change in the microbiome in response to disease state. Our analysis used statistically appropriate methods, which accounted for analytical challenges in describing the microbiome, decreasing the possibility of false positives, especially among the identified taxa (36,54). Our analysis has also addressed confounders, which may affect the microbiome and survival, including the strong individual microbiome signature.

However, our study has some limitations. First, our results focus on late-stage cancer patients in northern Europe, and therefore may not be broadly generalizable. There are reports of differences in the tumor microbiome between early and late stage CRC patients (55), and differences in the healthy microbiome between countries (56). However, past work has suggested that CRC is characterized by a set of organisms similar to the ones we identified, and our work overlaps with the results of a Chinese cohort, despite methodological differences (5,6,13). Additionally, we did not characterize our specific taxonomic profiles in a validation cohort, meaning the taxa separating tissue types and the tumor survival index may be specific features of our cohort, rather than able to predict survival in a broader population of late-stage CRC patients. Finally, we profiled the microbiome using 16S rRNA sequencing, with all the assumptions, benefits, and limitations of the technique. Our work is predicated on the assumption that phylogenetic similarity correlates to genetic and niche similarity. Without robust functional prediction and the ability to assemble genome units, we are limited in our mechanistic insight. However, our 16S sequencing is, in many cases, able to capture species or sub-species level resolution as the amplicon sequence variant ID, even if the name cannot be inferred accurately (57,58).

## CONCLUSION

We performed a nested case-control of the role of the microbiome in relapse free survival following primary resection in late-stage CRC patients. We identified clear differences in the microbiome between normal and tumor tissue and that a larger difference between tissue types was associated with poor prognosis. We found the tumor microbiome was associated with survival. This suggests a need to focus microbiome-based interventions at the tumor-specific community, rather trying to modify prognosis by changing the gut microbiome overall.

## Supporting information

Storms Checklist

## Data Availability

All data produced in the present study are available upon reasonable request to the authors

## DATA AVAILABILITY

Due to patient privacy concerns, sequences are available upon reasonable request. Patient data will not be released.

## ACKNOWLEDGEMENTS

The authors wish to thank the Department of Surgery, County Hospital Ryhov, for the collection of tissue biopsies. Thanks to the lab core at Centre for Translational Microbiome Research for support in extracting, processing, and sequencing the samples. We are also grateful to Cameron Martino, Marcus Fedarko, and Kalen Cantrell for their rapid responses to bug reports and feature requests for the gemelli and empress qiime2 plugins.

## AUTHOR CONTRIBUTIONS

RSO, MS, LE, and AM designed the study. RSO collected the tissue samples and reviewed medical records. JWD prepared the data. JWD performed the bioinformatic analysis; JWD and NB analyzed the data with advice from JTM. JWD drafted the manuscript. All authors reviewed and approved the final manuscript.

## Supplemental tables

Table S1. Patient characteristics in the cohort

Table S2. Per-subject predictors of the microbiome in CTF ordination space

Table S3. ASVs separating normal and tumor tissue based on differential ranking

Table S4. ASVs associated with the survival term in the interaction differential ranking model

Table S5. ASVs associated with the tissue term in the interaction differential ranking model

Table S6. ASVs associated with the interaction term in the interaction differential ranking model

Table S7. ASVs used for the tumor rPCA ALR calculation

## Supplemental Figures

Figure S1. A family level view of the microbiome

Figure S2. There is a strong individual effect on the microbiome

Figure S3. Metadata predictors of the microbiome in Compositional Tensor Factorization (CTF) space

Figure S4. Survival separates individuals by tumor-associated PCA quadrant

## Supplemental Files

File S1. Representative sequences separating tumor tissue

File S2. Representative sequences identified in the interaction model

File S3. Representative sequences for ASVs associated with PC1 in rPCA space

File S4. STORMS checklist

