## Supplemental Figures and Tables for "The local tumor microbiome is associated with survival in late-stage colorectal cancer patients"

**Supplemental tables**

Table S1. Patient characteristics in the cohort
Table S2. Per-Subject Predictors of the Microbiome in CTF ordination space
Table S3. ASVs separating tumor and normal tissue based on differential ranking
Table S4. Difference between normal and tumor tissue and survival
Table S5. Feature Ranks
Table S6. Feature Ranks for
Table S7. Tumor rPCA PC 1 features-associated ASVS for the ALR calculation

**Supplemental Figures**

Figure S1. A family level view of the microbiome
Figure S2. There is a strong individual effect on the microbiome
Figure S3. Metadata predictors of the microbiome in Compositional Tensor Factorization.
Figure S4. Survival separates individuals by tumor-associated rPCA quadrant

**Supplemental Files**
File S1. Representative sequences seperating tumor tissue
File S2. Representative sequences identified in the interaction model
File S3. Representative sequences for ASVs associated with PC 1 and PC 2 in rPCA space
File S4. STORMS Checklist

**Table S1. Patient characteristics in the cohort**

| | Short-term survival (< 2 years) (n=50) | Long-term survival (≥5 years) (n=51) | Total (n=101) |
|---|---|---|---|
| Patient characteristics at time of surgery | | | |
| **Age, years** | | | |
| *<60* | 6 (12.0) | 9 (17.7) | 15 (14.9) |
| *60-69* | 11 (22.0) | 19 (37.3) | 30 (29.7) |
| *70-74* | 16 (32.0) | 10 (19.6) | 26 (25.7) |
| *≥75* | 17 (34.0) | 13 (25.5) | 30 (29.7) |
| **Sex** | | | |
| *Female* | 24 (48.0) | 21 (41.2) | 45 (44.6) |
| *Male* | 26 (52.0) | 30 (58.8) | 56 (55.5) |
| **ASA score** | | | |
| *I (healthy)* | 14 (28.0) | 15 (29.4) | 29 (28.7) |
| *II (mild)* | 21 (42.0) | 28 (54.9) | 49 (48.5) |
| *III-IV (severe or worse)* | 15 (30.0) | 8 (15.7) | 23 (22.8) |
| **Pre-operative treatment** | | | |
| *None* | 41 (82.0) | 35 (68.6) | 76 (75.3) |
| *Radiotherapy* | 9 (18.0) | 16 (31.4) | 25 (24.7) |
| Tumor characteristics | | | |
| **Localization** | | | |
| Colon right | 18 (36.0) | 13 (25.5) | 31 (30.7) |
| Colon left | 11 (22.0) | 17 (33.3) | 28 (27.7) |
| Rectum | 21 (42.0) | 21 (41.2) | 42 (41.6) |
| **Mucinous cancer** | | | |
| no | 45 (90.0) | 45 (88.2) | 90 (89.1) |
| yes | 5 (10.0) | 6 (11.8) | 11 (10.9) |
| **TNM stage** | | | |
| III | 23 (46.0) | 46 (90.2) | 69 (68.3) |
| IV | 27 (54.0) | 5 (9.8) | 32 (31.7) |
| **Grade of differentiation** | | | |
| low | 22 (44.0) | 7 (13.7) | 29 (28.7) |
| medium | 24 (48.0) | 38 (74.5) | 62 (61.4) |
| high | 4 (8.0) | 6 (11.8) | 10 (9.9) |
| Surgical characteristics | | | |
| **Period of surgery** | | | |
| *1997-2005* | 19 (38.0) | 13 (25.5) | 32 (31.7) |
| *2006-2010* | 18 (36.0) | 23 (45.1) | 41 (40.6) |
| *2011-2017* | 13 (26.0) | 15 (29.4) | 28 (27.7) |
| **Radical operation** | | | |
| *no* | 14 (28.0) | 1 (2.0) | 15 (14.9) |
| *yes* | 36 (72.0) | 50 (98.0) | 86 (85.2) |
| **Microscopic radical operation** | | | |
| *no* | 13 (26.9) | 1 (2.0) | 14 (13.9) |
| *yes* | 37 (74.0) | 50 (98.0) | 87 (86.1) |

**Table S2. Per-Subject Predictors of the Microbiome in CTF ordination space**

| Predictor | Variation explained (%) | Crude p-value (999 permutations) |
|---|---|---|
| Age, years | 0.020 | 0.776 |
| Sex | 0.011 | 0.364 |
| ASA score | 0.034 | 0.140 |
| Localization | 0.011 | 0.778 |
| TNM Stage | 0.004 | 0.764 |
| Grade of differentiation | 0.007 | 0.931 |
| Period of Surgery | 0.151 | 0.001 |
| Survival | 0.020 | 0.113 |

**Table S3. ASVs separating tumor and normal tissue based on differential ranking**

| | | ASV ID | Prevalence | | Taxonomy |
|---|---|---|---|---|---|
| | | | ASV | Cumulative | |
| Normal tissue associated | 1 | Clos-b4ed8ec | 27.7% | 27.7% | p. Firmicutes; c. Clostridia; o. Clostridiales; f. Clostridiaceae 1; g. Clostridium sensu stricto 1 |
| | 2 | Lach-e23e6e7 | 21.8% | 39.6% | p. Firmicutes; c. Clostridia; o. Clostridiales; f. Lachnospiraceae; g. uncl f. Lachnospiraceae |
| | 3 | Bact-7ce20dc | 19.8% | 50.0% | p. Bacteroidetes; c. Bacteroidia; o. Bacteroidales; f. Bacteroidaceae; g. Bacteroides |
| | 4 | Marv-c55b486 | 36.1% | 68.8% | p. Firmicutes; c. Clostridia; o. Clostridiales; f. Lachnospiraceae; g. Marvinbryantia |
| | 5 | Bact-47d7490 | 18.8% | 73.3% | p. Bacteroidetes; c. Bacteroidia; o. Bacteroidales; f. Bacteroidaceae; g. Bacteroides |
| | 6 | Pept-1d2b00f | 19.8% | 74.8% | p. Firmicutes; c. Clostridia; o. Clostridiales; f. Peptostreptococcaceae; g. Peptostreptococcus |
| | 7 | Para-3d48874 | 21.3% | 79.7% | p. Bacteroidetes; c. Bacteroidia; o. Bacteroidales; f. Prevotellaceae; g. Paraprevotella |
| | 8 | Rumi-0f06bc7 | 24.8% | 82.2% | p. Firmicutes; c. Clostridia; o. Clostridiales; f. Ruminococcaceae; g. Ruminococcaceae UCG-002 |
| | 9 | Lach-11902be | 38.1% | 89.6% | p. Firmicutes; c. Clostridia; o. Clostridiales; f. Lachnospiraceae; g. uncl f. Lachnospiraceae |
| | 10 | Rumi-3efbc4a | 29.2% | 90.6% | p. Firmicutes; c. Clostridia; o. Clostridiales; f. Ruminococcaceae; g. uncl f. Ruminococcaceae |
| | 11 | Alis-520887a | 30.7% | 92.1% | p. Bacteroidetes; c. Bacteroidia; o. Bacteroidales; f. Rikenellaceae; g. Alistipes |
| | 12 | Bact-87e7824 | 15.3% | 93.1% | p. Bacteroidetes; c. Bacteroidia; o. Bacteroidales; f. Bacteroidaceae; g. Bacteroides |
| | 13 | Buty-da8b26f | 43.6% | 96.0% | p. Firmicutes; c. Clostridia; o. Clostridiales; f. Ruminococcaceae; g. Butyricicoccus |
| | 14 | Anae-4f9106b | 30.7% | 97.0% | p. Firmicutes; c. Clostridia; o. Clostridiales; f. Ruminococcaceae; g. Anaerotruncus |
| | 15 | Rumi-494e655 | 33.7% | 97.0% | p. Firmicutes; c. Clostridia; o. Clostridiales; f. Ruminococcaceae; g. Ruminococcaceae UCG-002 |
| | 16 | Anae-2ee585c | 26.2% | 97.5% | p. Firmicutes; c. Clostridia; o. Clostridiales; f. Lachnospiraceae; g. Anaerostipes |
| | 17 | Blau-400ef32 | 64.4% | 98.0% | p. Firmicutes; c. Clostridia; o. Clostridiales; f. Lachnospiraceae; g. Blautia |
| | 18 | Bifi-38b7580 | 41.6% | 98.0% | p. Actinobacteria; c. Actinobacteria; o. Bifidobacteriales; f. Bifidobacteriaceae; g. Bifidobacterium |
| | 19 | Rumi-a39d801 | 19.3% | 99.0% | p. Firmicutes; c. Clostridia; o. Clostridiales; f. Ruminococcaceae; g. Ruminococcus 2 |
| | 20 | Rumi-75a7dd0 | 24.8% | 99.0% | p. Firmicutes; c. Clostridia; o. Clostridiales; f. Ruminococcaceae; g. Ruminococcus 2 |
| | 21 | Rumi-f7d40e3 | 20.3% | 99.5% | p. Firmicutes; c. Clostridia; o. Clostridiales; f. Ruminococcaceae; g. Ruminococcus 2 |
| | 22 | Blau-8f6e2a9 | 95.0% | 100.0% | p. Firmicutes; c. Clostridia; o. Clostridiales; f. Lachnospiraceae; g. Blautia |
| Tumor Tissue Associated | 1 | Fuso-e47b7c5 | 25.2% | 25.2% | p. Fusobacteria; c. Fusobacteriia; o. Fusobacteriales; f. Fusobacteriaceae; g. Fusobacterium |
| | 2 | Camp-5b14d87 | 27.2% | 46.5% | p. Proteobacteria; c. Epsilonproteobacteria; o. Campylobacterales; f. Campylobacteraceae; g. Campylobacter |
| | 3 | Fuso-d2f4ba7 | 12.4% | 50.0% | p. Fusobacteria; c. Fusobacteriia; o. Fusobacteriales; f. Fusobacteriaceae; g. Fusobacterium |
| | 4 | Hung-78c2b8e | 33.7% | 61.9% | p. Firmicutes; c. Clostridia; o. Clostridiales; f. Lachnospiraceae; g. Hungatella |
| | 5 | Fuso-7fa9543 | 21.3% | 66.8% | p. Fusobacteria; c. Fusobacteriia; o. Fusobacteriales; f. Fusobacteriaceae; g. Fusobacterium |
| | 6 | Gran-a5b69f1 | 42.6% | 79.7% | p. Firmicutes; c. Bacilli; o. Lactobacillales; f. Carnobacteriaceae; g. Granulicatella |
| | 7 | Bact-26547c9 | 34.2% | 87.6% | p. Bacteroidetes; c. Bacteroidia; o. Bacteroidales; f. Bacteroidaceae; g. Bacteroides |
| | 8 | Stre-d3829c5 | 19.3% | 88.1% | p. Firmicutes; c. Bacilli; o. Lactobacillales; f. Streptococcaceae; g. Streptococcus |
| | 9 | Mass-bfb5b51 | 38.1% | 91.6% | p. Proteobacteria; c. Betaproteobacteria; o. Burkholderiales; f. Oxalobacteraceae; g. Massilia |
| | 10 | Para-8d83490 | 11.9% | 92.6% | p. Bacteroidetes; c. Bacteroidia; o. Bacteroidales; f. Porphyromonadaceae; g. Parabacteroides |
| | 11 | Porp-69023d0 | 18.8% | 92.6% | p. Bacteroidetes; c. Bacteroidia; o. Bacteroidales; f. Porphyromonadaceae; g. Porphyromonas |
| | 12 | Lach-7cd4efa | 54.0% | 95.5% | p. Firmicutes; c. Clostridia; o. Clostridiales; f. Lachnospiraceae; g. Lachnoclostridium |
| | 13 | Hung-82c448c | 29.7% | 96.0% | p. Firmicutes; c. Clostridia; o. Clostridiales; f. Lachnospiraceae; g. Hungatella |
| | 14 | Para-98a6938 | 17.3% | 97.5% | p. Proteobacteria; c. Betaproteobacteria; o. Burkholderiales; f. Alcaligenaceae; g. Parasutterella |
| | 15 | Stre-101637a | 36.6% | 98.5% | p. Firmicutes; c. Bacilli; o. Lactobacillales; f. Streptococcaceae; g. Streptococcus |
| | 16 | Sutt-1d5d65b | 19.3% | 99.5% | p. Proteobacteria; c. Betaproteobacteria; o. Burkholderiales; f. Alcaligenaceae; g. Sutterella |
| | 17 | Rumi-eee1408 | 20.3% | 100.0% | p. Firmicutes; c. Clostridia; o. Clostridiales; f. Ruminococcaceae; g. Ruminococcaceae UCG-014 |

**Table S4. Difference between normal and tumor tissue and survival**

| Metric | Crude | Fully adjusted[1] |
|---|---|---|
| Binary Jaccard (0.1 unit) | 1.53 (1.00, 2.34) | 1.61 (0.83, 3.15) |
| Aitchison (10 units) | 1.91 (1.12, 3.27) | 1.42 (0.59, 3.44) |
| CTF (1 unit) | 1.20 (0.54, 2.67) | 1.94 (0.54, 6.98) |
| Bray Curtis (0.1 unit) | 1.63 (1.18, 2.26) | 1.70 (1.06, 2.73) |
| unweighted UniFrac (0.1 unit) | 1.91 (1.06, 3.43) | 2.01 (0.80, 5.02) |
| weighted UniFrac (0.1 unit) | 1.36 (1.11, 1.66) | 1.48 (1.08, 2.03) |

[1]Adjusted for age, sex, ASA score, tumor localization, TNM stage, differentiation grade, surgery period, and radical surgery

**Table S5. Feature Ranks for tumor vs normal tissue, based on long survival**

| | | Feature-id | Prevalence | | Taxonomy |
|---|---|---|---|---|---|
| | | | ASV | Cumulative | |
| Normal tissue associated | 1 | Bact-47d7490 | 18.8% | 18.8% | p. Bacteroidetes; c. Bacteroidia; o. Bacteroidales; f. Bacteroidaceae; g. Bacteroides |
| | 2 | Pept-1d2b00f | 19.8% | 34.2% | p. Firmicutes; c. Clostridia; o. Clostridiales; f. Peptostreptococcaceae; g. Peptostreptococcus |
| | 3 | Bact-208134d | 17.8% | 45.5% | p. Bacteroidetes; c. Bacteroidia; o. Bacteroidales; f. Bacteroidaceae; g. Bacteroides |
| | 4 | Para-3d48874 | 21.3% | 52.5% | p. Bacteroidetes; c. Bacteroidia; o. Bacteroidales; f. Prevotellaceae; g. Paraprevotella |
| | 5 | Bact-5859c64 | 25.2% | 64.4% | p. Bacteroidetes; c. Bacteroidia; o. Bacteroidales; f. Bacteroidaceae; g. Bacteroides |
| | 6 | Barn-f674778 | 19.3% | 70.8% | p. Bacteroidetes; c. Bacteroidia; o. Bacteroidales; f. Porphyromonadaceae; g. Barnesiella |
| | 7 | Marv-c55b486 | 36.1% | 80.2% | p. Firmicutes; c. Clostridia; o. Clostridiales; f. Lachnospiraceae; g. Marvinbryantia |
| | 8 | Clos-b4ed8ec | 27.7% | 84.2% | p. Firmicutes; c. Clostridia; o. Clostridiales; f. Clostridiaceae 1; g. Clostridium sensu stricto 1 |
| | 9 | Para-218f40f | 35.6% | 89.6% | p. Bacteroidetes; c. Bacteroidia; o. Bacteroidales; f. Porphyromonadaceae; g. Parabacteroides |
| | 10 | Bifi-38b7580 | 41.6% | 92.1% | p. Actinobacteria; c. Actinobacteria; o. Bifidobacteriales; f. Bifidobacteriaceae; g. Bifidobacterium |
| | 11 | Desu-fa6956a | 23.3% | 93.1% | p. Proteobacteria; c. Deltaproteobacteria; o. Desulfovibrionales; f. Desulfovibrionaceae; g. Desulfovibrio |
| | 12 | Meth-26639d9 | 24.8% | 93.1% | p. Euryarchaeota; c. Methanobacteria; o. Methanobacteriales; f. Methanobacteriaceae; g. Methanobrevibacter |
| | 13 | Lach-e23e6e7 | 21.8% | 93.1% | p. Firmicutes; c. Clostridia; o. Clostridiales; f. Lachnospiraceae; g. unsp. f. Lachnospiraceae |
| | 14 | Bact-b74ab69 | 19.3% | 94.1% | p. Bacteroidetes; c. Bacteroidia; o. Bacteroidales; f. Bacteroidaceae; g. Bacteroides |
| | 15 | Stre-a4cd615 | 16.8% | 95.5% | p. Firmicutes; c. Bacilli; o. Lactobacillales; f. Streptococcaceae; g. Streptococcus |
| | 16 | Sutt-381ef54 | 19.8% | 96.0% | p. Proteobacteria; c. Betaproteobacteria; o. Burkholderiales; f. Alcaligenaceae; g. Sutterella |
| | 17 | Bact-7ce20dc | 19.8% | 96.0% | p. Bacteroidetes; c. Bacteroidia; o. Bacteroidales; f. Bacteroidaceae; g. Bacteroides |
| | 18 | Buty-da8b26f | 43.6% | 97.0% | p. Firmicutes; c. Clostridia; o. Clostridiales; f. Ruminococcaceae; g. Butyricicoccus |
| | 19 | Rumi-4a1547a | 46.5% | 97.0% | p. Firmicutes; c. Clostridia; o. Clostridiales; f. Ruminococcaceae; g. Ruminiclostridium 6 |
| | 20 | Rumi-8dc9986 | 21.8% | 98.0% | p. Firmicutes; c. Clostridia; o. Clostridiales; f. Ruminococcaceae; g. Ruminococcaceae UCG-002 |
| | 21 | Bifi-f633cf9 | 24.8% | 98.0% | p. Actinobacteria; c. Actinobacteria; o. Bifidobacteriales; f. Bifidobacteriaceae; g. Bifidobacterium |
| | 22 | Turi-c96cf26 | 25.2% | 98.0% | p. Firmicutes; c. Erysipelotrichia; o. Erysipelotrichales; f. Erysipelotrichaceae; g. Turicibacter |
| | 23 | Mass-cabd729 | 24.8% | 99.0% | p. Proteobacteria; c. Betaproteobacteria; o. Burkholderiales; f. Oxalobacteraceae; g. Massilia |
| | 24 | Rumi-75a7dd0 | 24.8% | 99.0% | p. Firmicutes; c. Clostridia; o. Clostridiales; f. Ruminococcaceae; g. Ruminococcus 2 |
| | 25 | Hold-5637d39 | 14.4% | 99.0% | p. Firmicutes; c. Erysipelotrichia; o. Erysipelotrichales; f. Erysipelotrichaceae; g. Holdemanella |
| | 26 | Haem-18eb929 | 41.1% | 99.5% | p. Proteobacteria; c. Gammaproteobacteria; o. Pasteurellales; f. Pasteurellaceae; g. Haemophilus |
| | 27 | Chri-b03de84 | 43.1% | 100.0% | p. Firmicutes; c. Clostridia; o. Clostridiales; f. Christensenellaceae; g. Christensenellaceae R-7 group |

| | | Feature-id | Prevalence | | Taxonomy |
|---|---|---|---|---|---|
| | | | ASV | Cumulative | |
| Tumor tissue | 1 | Hung-78c2b8e | 33.7% | 33.7% | p. Firmicutes; c. Clostridia; o. Clostridiales; f. Lachnospiraceae; g. Hungatella |
| | 2 | Fuso-e47b7c5 | 25.2% | 47.0% | p. Fusobacteria; c. Fusobacteriia; o. Fusobacteriales; f. Fusobacteriaceae; g. Fusobacterium |
| | 3 | Lach-7cd4efa | 54.0% | 66.8% | p. Firmicutes; c. Clostridia; o. Clostridiales; f. Lachnospiraceae; g. Lachnoclostridium |
| | 4 | Camp-5b14d87 | 27.2% | 77.7% | p. Proteobacteria; c. Epsilonproteobacteria; o. Campylobacterales; f. Campylobacteraceae; g. Campylobacter |
| | 5 | Rose-3a89328 | 37.1% | 86.1% | p. Firmicutes; c. Clostridia; o. Clostridiales; f. Lachnospiraceae; g. Roseburia |
| | 6 | Sutt-1d5d65b | 19.3% | 88.6% | p. Proteobacteria; c. Betaproteobacteria; o. Burkholderiales; f. Alcaligenaceae; g. Sutterella |
| | 7 | Stre-d3829c5 | 19.3% | 91.1% | p. Firmicutes; c. Bacilli; o. Lactobacillales; f. Streptococcaceae; g. Streptococcus |
| | 8 | Buty-62c3369 | 25.2% | 92.1% | p. Firmicutes; c. Clostridia; o. Clostridiales; f. Ruminococcaceae; g. Butyricicoccus |
| | 9 | Hung-82c448c | 29.7% | 93.6% | p. Firmicutes; c. Clostridia; o. Clostridiales; f. Lachnospiraceae; g. Hungatella |
| | 10 | Fuso-d2f4ba7 | 12.4% | 93.6% | p. Fusobacteria; c. Fusobacteriia; o. Fusobacteriales; f. Fusobacteriaceae; g. Fusobacterium |
| | 11 | Mass-bfb5b51 | 38.1% | 95.5% | p. Proteobacteria; c. Betaproteobacteria; o. Burkholderiales; f. Oxalobacteraceae; g. Massilia |
| | 12 | Bact-866ea1c | 13.9% | 95.5% | p. Bacteroidetes; c. Bacteroidia; o. Bacteroidales; f. Bacteroidaceae; g. Bacteroides |
| | 13 | Porp-0799969 | 21.3% | 95.5% | p. Bacteroidetes; c. Bacteroidia; o. Bacteroidales; f. Porphyromonadaceae; g. Porphyromonas |
| | 14 | Dial-08dda4f | 13.4% | 96.0% | p. Firmicutes; c. Negativicutes; o. Selenomonadales; f. Veillonellaceae; g. Dialister |
| | 15 | Barn-53f976e | 14.9% | 97.5% | p. Bacteroidetes; c. Bacteroidia; o. Bacteroidales; f. Porphyromonadaceae; g. Barnesiella |
| | 16 | Bact-0246885 | 20.8% | 97.5% | p. Bacteroidetes; c. Bacteroidia; o. Bacteroidales; f. Bacteroidaceae; g. Bacteroides |
| | 17 | Phas-fb7a1c0 | 20.8% | 97.5% | p. Firmicutes; c. Negativicutes; o. Selenomonadales; f. Acidaminococcaceae; g. Phascolarctobacterium |
| | 18 | Rumi-6a57339 | 31.2% | 98.0% | p. Firmicutes; c. Clostridia; o. Clostridiales; f. Ruminococcaceae; g. unsp. f. Ruminococcaceae |
| | 19 | Esch-ffc36e2 | 90.6% | 100.0% | p. Proteobacteria; c. Gammaproteobacteria; o. Enterobacteriales; f. Enterobacteriaceae; g. Escherichia/Shigella |

**Table S6. Feature Ranks for interaction term (short survival in tumor tissue) from**

| | | Feature-id | Prevalence | | Taxonomy |
|---|---|---|---|---|---|
| | | | ASV | Cumulative | |
| Normal Tissue | 1 | Chri-fb2e43a | 20.3% | 20.3% | p. Firmicutes; c. Clostridia; o. Clostridiales; f. Christensenellaceae; g. Christensenellaceae R-7 group |
| | 2 | Anae-2ee585c | 26.2% | 40.6% | p. Firmicutes; c. Clostridia; o. Clostridiales; f. Lachnospiraceae; g. Anaerostipes |
| | 3 | Alis-0ce6000 | 14.9% | 50.5% | p. Bacteroidetes; c. Bacteroidia; o. Bacteroidales; f. Rikenellaceae; g. Alistipes |
| | 4 | Barn-53f976e | 14.9% | 55.9% | p. Bacteroidetes; c. Bacteroidia; o. Bacteroidales; f. Porphyromonadaceae; g. Barnesiella |
| | 5 | Chri-f7035cb | 20.8% | 63.9% | p. Firmicutes; c. Clostridia; o. Clostridiales; f. Christensenellaceae; g. Christensenellaceae R-7 group |
| | 6 | Romb-df3d611 | 55.0% | 78.7% | p. Firmicutes; c. Clostridia; o. Clostridiales; f. Peptostreptococcaceae; g. Romboutsia |
| | 7 | Rumi-6c7b500 | 19.3% | 80.7% | p. Firmicutes; c. Clostridia; o. Clostridiales; f. Ruminococcaceae; g. Ruminococcaceae NK4A214 group |
| | 8 | Rhod-1667bf7 | 24.3% | 83.2% | p. Proteobacteria; c. Alphaproteobacteria; o. Rhodospirillales; f. Rhodospirillaceae; g. unsp. f. Rhodospirillaceae |
| | 9 | Lach-7079408 | 50.5% | 88.1% | p. Firmicutes; c. Clostridia; o. Clostridiales; f. Lachnospiraceae; g. Lachnospiraceae NK4A136 group |
| | 10 | Rumi-6a57339 | 31.2% | 91.6% | p. Firmicutes; c. Clostridia; o. Clostridiales; f. Ruminococcaceae; g. unsp. f. Ruminococcaceae |
| | 11 | Alis-520887a | 30.7% | 93.6% | p. Bacteroidetes; c. Bacteroidia; o. Bacteroidales; f. Rikenellaceae; g. Alistipes |
| | 12 | Sene-781430c | 24.8% | 93.6% | p. Actinobacteria; c. Coriobacteriia; o. Coriobacteriales; f. Coriobacteriaceae; g. Senegalimassilia |
| | 13 | Rumi-3efbc4a | 29.2% | 93.6% | p. Firmicutes; c. Clostridia; o. Clostridiales; f. Ruminococcaceae; g. unsp. f. Ruminococcaceae |
| | 14 | Buty-62c3369 | 25.2% | 95.0% | p. Firmicutes; c. Clostridia; o. Clostridiales; f. Ruminococcaceae; g. Butyricicoccus |
| | 15 | Lach-7cd4efa | 54.0% | 97.0% | p. Firmicutes; c. Clostridia; o. Clostridiales; f. Lachnospiraceae; g. Lachnoclostridium |
| | 16 | Rose-3a89328 | 37.1% | 99.0% | p. Firmicutes; c. Clostridia; o. Clostridiales; f. Lachnospiraceae; g. Roseburia |
| | 17 | Rose-d3864fc | 34.2% | 99.0% | p. Firmicutes; c. Clostridia; o. Clostridiales; f. Lachnospiraceae; g. Roseburia |
| | 18 | Lach-3a2e845 | 33.7% | 99.5% | p. Firmicutes; c. Clostridia; o. Clostridiales; f. Lachnospiraceae; g. unsp. f. Lachnospiraceae |
| | 19 | Hung-78c2b8e | 33.7% | 100.0% | p. Firmicutes; c. Clostridia; o. Clostridiales; f. Lachnospiraceae; g. Hungatella |

| | | Feature-id | Prevalence | | Taxonomy |
|---|---|---|---|---|---|
| | | | ASV | Cumulative | |
| Tumor Tissue | 1 | Pept-1d2b00f | 19.8% | 19.8% | p. Firmicutes; c. Clostridia; o. Clostridiales; f. Peptostreptococcaceae; g. Peptostreptococcus |
| | 2 | Bact-47d7490 | 18.8% | 34.2% | p. Bacteroidetes; c. Bacteroidia; o. Bacteroidales; f. Bacteroidaceae; g. Bacteroides |
| | 3 | Bifi-c299192 | 14.4% | 45.0% | p. Actinobacteria; c. Actinobacteria; o. Bifidobacteriales; f. Bifidobacteriaceae; g. Bifidobacterium |
| | 4 | Bact-208134d | 17.8% | 55.9% | p. Bacteroidetes; c. Bacteroidia; o. Bacteroidales; f. Bacteroidaceae; g. Bacteroides |
| | 5 | Sutt-381ef54 | 19.8% | 67.3% | p. Proteobacteria; c. Betaproteobacteria; o. Burkholderiales; f. Alcaligenaceae; g. Sutterella |
| | 6 | Para-3d48874 | 21.3% | 71.3% | p. Bacteroidetes; c. Bacteroidia; o. Bacteroidales; f. Prevotellaceae; g. Paraprevotella |
| | 7 | Bact-5859c64 | 25.2% | 77.2% | p. Bacteroidetes; c. Bacteroidia; o. Bacteroidales; f. Bacteroidaceae; g. Bacteroides |
| | 8 | Veil-24522f7 | 27.7% | 81.7% | p. Firmicutes; c. Negativicutes; o. Selenomonadales; f. Veillonellaceae; g. Veillonella |
| | 9 | Barn-f674778 | 19.3% | 85.1% | p. Bacteroidetes; c. Bacteroidia; o. Bacteroidales; f. Porphyromonadaceae; g. Barnesiella |
| | 10 | Bact-26547c9 | 34.2% | 92.1% | p. Bacteroidetes; c. Bacteroidia; o. Bacteroidales; f. Bacteroidaceae; g. Bacteroides |
| | 11 | Chri-b03de84 | 43.1% | 95.0% | p. Firmicutes; c. Clostridia; o. Clostridiales; f. Christensenellaceae; g. Christensenellaceae R-7 group |
| | 12 | Bact-98fcc7e | 53.5% | 96.0% | p. Bacteroidetes; c. Bacteroidia; o. Bacteroidales; f. Bacteroidaceae; g. Bacteroides |
| | 13 | Fuso-7fa9543 | 21.3% | 96.0% | p. Fusobacteria; c. Fusobacteriia; o. Fusobacteriales; f. Fusobacteriaceae; g. Fusobacterium |
| | 14 | Sutt-222e21f | 25.2% | 97.0% | p. Proteobacteria; c. Betaproteobacteria; o. Burkholderiales; f. Alcaligenaceae; g. Sutterella |
| | 15 | Para-218f40f | 35.6% | 97.5% | p. Bacteroidetes; c. Bacteroidia; o. Bacteroidales; f. Porphyromonadaceae; g. Parabacteroides |
| | 16 | Bifi-f633cf9 | 24.8% | 97.5% | p. Actinobacteria; c. Actinobacteria; o. Bifidobacteriales; f. Bifidobacteriaceae; g. Bifidobacterium |
| | 17 | Clos-875f9ef | 27.2% | 99.0% | p. Firmicutes; c. Clostridia; o. Clostridiales; f. Clostridiaceae 1; g. Clostridium sensu stricto 1 |
| | 18 | Rose-d91b5c7 | 34.2% | 99.5% | p. Firmicutes; c. Clostridia; o. Clostridiales; f. Lachnospiraceae; g. Roseburia |
| | 19 | Fret-2b8622a | 22.3% | 99.5% | p. Synergistetes; c. Synergistia; o. Synergistales; f. Synergistaceae; g. Fretibacterium |
| | 20 | Fuso-62b2f8b | 19.3% | 99.5% | p. Fusobacteria; c. Fusobacteriia; o. Fusobacteriales; f. Fusobacteriaceae; g. Fusobacterium |
| | 21 | Sutt-dfc8256 | 33.7% | 99.5% | p. Proteobacteria; c. Betaproteobacteria; o. Burkholderiales; f. Alcaligenaceae; g. Sutterella |
| | 22 | Marv-c55b486 | 36.1% | 99.5% | p. Firmicutes; c. Clostridia; o. Clostridiales; f. Lachnospiraceae; g. Marvinbryantia |
| | 23 | Rumi-4a1547a | 46.5% | 99.5% | p. Firmicutes; c. Clostridia; o. Clostridiales; f. Ruminococcaceae; g. Ruminiclostridium 6 |
| | 24 | Bact-b74ab69 | 19.3% | 99.5% | p. Bacteroidetes; c. Bacteroidia; o. Bacteroidales; f. Bacteroidaceae; g. Bacteroides |
| | 25 | Gast-90e9bc6 | 24.8% | 99.5% | p. Cyanobacteria; c. Melainabacteria; o. Gastranaerophilales; f. unsp. o. Gastranaerophilales; g. unsp. o. Gastranaerophilales |
| | 26 | Meth-26639d9 | 24.8% | 99.5% | p. Euryarchaeota; c. Methanobacteria; o. Methanobacteriales; f. Methanobacteriaceae; g. Methanobrevibacter |
| | 27 | Dial-35fd415 | 60.4% | 99.5% | p. Firmicutes; c. Negativicutes; o. Selenomonadales; f. Veillonellaceae; g. Dialister |
| | 28 | Lach-3765e25 | 30.2% | 99.5% | p. Firmicutes; c. Clostridia; o. Clostridiales; f. Lachnospiraceae; g. Lachnospiraceae NK4A136 group |
| | 29 | Desu-fa6956a | 23.3% | 99.5% | p. Proteobacteria; c. Deltaproteobacteria; o. Desulfovibrionales; f. Desulfovibrionaceae; g. Desulfovibrio |
| | 30 | Rumi-8e121f6 | 38.1% | 100.0% | p. Firmicutes; c. Clostridia; o. Clostridiales; f. Ruminococcaceae; g. unsp. f. Ruminococcaceae |

### Table S7. Tumor rPCA PC 1 features-associated ASVS for the ALR calculation

| Group | Feature-id | Taxonomy |
|---|---|---|
| Long-term Survival | Bact-70d55ba | p. Bacteroidetes; c. Bacteroidia; o. Bacteroidales; f. Bacteroidaceae; g. Bacteroides |
| | Bact-b6635d6 | p. Bacteroidetes; c. Bacteroidia; o. Bacteroidales; f. Bacteroidaceae; g. Bacteroides |
| | Faec-4516aa6 | p. Firmicutes; c. Clostridia; o. Clostridiales; f. Ruminococcaceae; g. Faecalibacterium |
| | Faec-c728ad6 | p. Firmicutes; c. Clostridia; o. Clostridiales; f. Ruminococcaceae; g. Faecalibacterium |
| | Faec-22f4ee9 | p. Firmicutes; c. Clostridia; o. Clostridiales; f. Ruminococcaceae; g. Faecalibacterium |
| | Subd-c6fdf63 | p. Firmicutes; c. Clostridia; o. Clostridiales; f. Ruminococcaceae; g. Subdoligranulum |
| | Rose-b264ac8 | p. Firmicutes; c. Clostridia; o. Clostridiales; f. Lachnospiraceae; g. Roseburia |
| | Subd-b6a1cfc | p. Firmicutes; c. Clostridia; o. Clostridiales; f. Ruminococcaceae; g. Subdoligranulum |
| | Fusi-9df2517 | p. Firmicutes; c. Clostridia; o. Clostridiales; f. Lachnospiraceae; g. Fusicatenibacter |
| | Bifi-3f3a0ea | p. Actinobacteria; c. Actinobacteria; o. Bifidobacteriales; f. Bifidobacteriaceae; g. Bifidobacterium |
| Short-term Survival | Pept-f148c98 | p. Firmicutes; c. Clostridia; o. Clostridiales; f. Peptostreptococcaceae; g. Peptostreptococcus |
| | Bact-47f3d64 | p. Bacteroidetes; c. Bacteroidia; o. Bacteroidales; f. Bacteroidaceae; g. Bacteroides |
| | Dial-63719d6 | p. Firmicutes; c. Negativicutes; o. Selenomonadales; f. Veillonellaceae; g. Dialister |
| | Parv-8f3c8cb | p. Firmicutes; c. Clostridia; o. Clostridiales; f. Family XI; g. Parvimonas |
| | Fuso-e47b7c5 | p. Fusobacteria; c. Fusobacteriia; o. Fusobacteriales; f. Fusobacteriaceae; g. Fusobacterium |
| | Fret-2b8622a | p. Synergistetes; c. Synergistia; o. Synergistales; f. Synergistaceae; g. Fretibacterium |
| | Solo-114d0ae | p. Firmicutes; c. Erysipelotrichia; o. Erysipelotrichales; f. Erysipelotrichaceae; g. Solobacterium |
| | Clos-a82bde6 | p. Firmicutes; c. Clostridia; o. Clostridiales; o. Clostridiales; o. Clostridiales |
| | Camp-5b14d87 | p. Proteobacteria; c. Epsilonproteobacteria; o. Campylobacterales; f. Campylobacteraceae; g. Campylobacter |
| | Bact-26547c9 | p. Bacteroidetes; c. Bacteroidia; o. Bacteroidales; f. Bacteroidaceae; g. Bacteroides |
| | Gran-a5b69f1 | p. Firmicutes; c. Bacilli; o. Lactobacillales; f. Carnobacteriaceae; g. Granulicatella |
| | Lach-7273e6e | p. Firmicutes; c. Clostridia; o. Clostridiales; f. Lachnospiraceae; g. Lachnospiraceae FCS020 group |
| | Porp-69023d0 | p. Bacteroidetes; c. Bacteroidia; o. Bacteroidales; f. Porphyromonadaceae; g. Porphyromonas |
| | Fuso-7fa9543 | p. Fusobacteria; c. Fusobacteriia; o. Fusobacteriales; f. Fusobacteriaceae; g. Fusobacterium |
| | Geme-7bd174e | p. Firmicutes; c. Bacilli; o. Bacillales; f. Family XI; g. Gemella |
| | Lach-3682830 | p. Firmicutes; c. Clostridia; o. Clostridiales; f. Lachnospiraceae; g. Lachnospiraceae FCS020 group |
| | Bifi-37020b4 | p. Actinobacteria; c. Actinobacteria; o. Bifidobacteriales; f. Bifidobacteriaceae; g. Bifidobacterium |
| | Fuso-d2f4ba7 | p. Fusobacteria; c. Fusobacteriia; o. Fusobacteriales; f. Fusobacteriaceae; g. Fusobacterium |
| | Bact-605c17c | p. Bacteroidetes; c. Bacteroidia; o. Bacteroidales; f. Bacteroidaceae; g. Bacteroides |
| | Fret-fd4122f | p. Synergistetes; c. Synergistia; o. Synergistales; f. Synergistaceae; g. Fretibacterium |
| | Camp-7e4c8c9 | p. Proteobacteria; c. Epsilonproteobacteria; o. Campylobacterales; f. Campylobacteraceae; g. Campylobacter |
| | Porp-1693b47 | p. Bacteroidetes; c. Bacteroidia; o. Bacteroidales; f. Porphyromonadaceae; g. Porphyromonas |
| | Bifi-d1d2824 | p. Actinobacteria; c. Actinobacteria; o. Bifidobacteriales; f. Bifidobacteriaceae; g. Bifidobacterium |
| | Rumi-aaa27b7 | p. Firmicutes; c. Clostridia; o. Clostridiales; f. Ruminococcaceae; g. Ruminiclostridium 5 |
| | Rumi-af67155 | p. Firmicutes; c. Clostridia; o. Clostridiales; f. Ruminococcaceae; g. Ruminococcaceae UCG-005 |

**Table S 8. Tumor rPCA PC 2 associated features used in the ALR calculation**

| | ASV ID | Taxonomy |
|---|---|---|
| Long-term Survival | Osci-d425791 | p. Firmicutes; c. Clostridia; o. Clostridiales; f. Ruminococcaceae; g. Oscillibacter |
| | Bifi-d1d2824 | p. Actinobacteria; c. Actinobacteria; o. Bifidobacteriales; f. Bifidobacteriaceae; g. Bifidobacterium |
| | Bact-605c17c | p. Bacteroidetes; c. Bacteroidia; o. Bacteroidales; f. Bacteroidaceae; g. Bacteroides |
| | Lach-b61ab87 | p. Firmicutes; c. Clostridia; o. Clostridiales; f. Lachnospiraceae; f. Lachnospiraceae |
| | Bifi-37020b4 | p. Actinobacteria; c. Actinobacteria; o. Bifidobacteriales; f. Bifidobacteriaceae; g. Bifidobacterium |
| | Alis-0bf0743 | p. Bacteroidetes; c. Bacteroidia; o. Bacteroidales; f. Rikenellaceae; g. Alistipes |
| | Bact-b24ef5f | p. Bacteroidetes; c. Bacteroidia; o. Bacteroidales; f. Bacteroidaceae; g. Bacteroides |
| | Rose-f6bea81 | p. Firmicutes; c. Clostridia; o. Clostridiales; f. Lachnospiraceae; g. Roseburia |
| Short-term Survival | Bact-47f3d64 | p. Bacteroidetes; c. Bacteroidia; o. Bacteroidales; f. Bacteroidaceae; g. Bacteroides |
| | Bact-26547c9 | p. Bacteroidetes; c. Bacteroidia; o. Bacteroidales; f. Bacteroidaceae; g. Bacteroides |
| | Bact-4abaa48 | p. Bacteroidetes; c. Bacteroidia; o. Bacteroidales; f. Bacteroidaceae; g. Bacteroides |
| | Dial-35fd415 | p. Firmicutes; c. Negativicutes; o. Selenomonadales; f. Veillonellaceae; g. Dialister |
| | Esch-ffc36e2 | p. Proteobacteria; c. Gammaproteobacteria; o. Enterobacteriales; f. Enterobacteriaceae; g. Escherichia/Shigella |
| | Dial-63719d6 | p. Firmicutes; c. Negativicutes; o. Selenomonadales; f. Veillonellaceae; g. Dialister |
| | Camp-5b14d87 | p. Proteobacteria; c. Epsilonproteobacteria; o. Campylobacterales; f. Campylobacteraceae; g. Campylobacter |
| | Bact-b66635d6 | p. Bacteroidetes; c. Bacteroidia; o. Bacteroidales; f. Bacteroidaceae; g. Bacteroides |
| | Fuso-fb12967 | p. Fusobacteria; c. Fusobacteriia; o. Fusobacteriales; f. Fusobacteriaceae; g. Fusobacterium |
| | Bact-6251bd9 | p. Bacteroidetes; c. Bacteroidia; o. Bacteroidales; f. Bacteroidaceae; g. Bacteroides |
| | Pept-f148c98 | p. Firmicutes; c. Clostridia; o. Clostridiales; f. Peptostreptococcaceae; g. Peptostreptococcus |
| | Faec-4516aa6 | p. Firmicutes; c. Clostridia; o. Clostridiales; f. Ruminococcaceae; g. Faecalibacterium |
| | Bact-7440aa8 | p. Bacteroidetes; c. Bacteroidia; o. Bacteroidales; f. Bacteroidaceae; g. Bacteroides |
| | Fuso-e47b7c5 | p. Fusobacteria; c. Fusobacteriia; o. Fusobacteriales; f. Fusobacteriaceae; g. Fusobacterium |
| | Fuso-7fa9543 | p. Fusobacteria; c. Fusobacteriia; o. Fusobacteriales; f. Fusobacteriaceae; g. Fusobacterium |
| | Fuso-d2f4ba7 | p. Fusobacteria; c. Fusobacteriia; o. Fusobacteriales; f. Fusobacteriaceae; g. Fusobacterium |
| | Faec-bbae6ed | p. Firmicutes; c. Clostridia; o. Clostridiales; f. Ruminococcaceae; g. Faecalibacterium |
| | Lach-02e1e97 | p. Firmicutes; c. Clostridia; o. Clostridiales; f. Lachnospiraceae; g. Lachnospiraceae UCG-010 |
| | Porp-1693b47 | p. Bacteroidetes; c. Bacteroidia; o. Bacteroidales; f. Porphyromonadaceae; g. Porphyromonas |
| | Stre-d3829c5 | p. Firmicutes; c. Bacilli; o. Lactobacillales; f. Streptococcaceae; g. Streptococcus |
| | Porp-69023d0 | p. Bacteroidetes; c. Bacteroidia; o. Bacteroidales; f. Porphyromonadaceae; g. Porphyromonas |
| | Bact-70d55ba | p. Bacteroidetes; c. Bacteroidia; o. Bacteroidales; f. Bacteroidaceae; g. Bacteroides |
| | Fuso-62b2f8b | p. Fusobacteria; c. Fusobacteriia; o. Fusobacteriales; f. Fusobacteriaceae; g. Fusobacterium |
| | Parv-8f3c8cb | p. Firmicutes; c. Clostridia; o. Clostridiales; f. Family XI; g. Parvimonas |
| | Faec-c728ad6 | p. Firmicutes; c. Clostridia; o. Clostridiales; f. Ruminococcaceae; g. Faecalibacterium |
| | Fret-2b8622a | p. Synergistetes; c. Synergistia; o. Synergistales; f. Synergistaceae; g. Fretibacterium |
| | Dial-08dda4f | p. Firmicutes; c. Negativicutes; o. Selenomonadales; f. Veillonellaceae; g. Dialister |
| | Fret-fd4122f | p. Synergistetes; c. Synergistia; o. Synergistales; f. Synergistaceae; g. Fretibacterium |

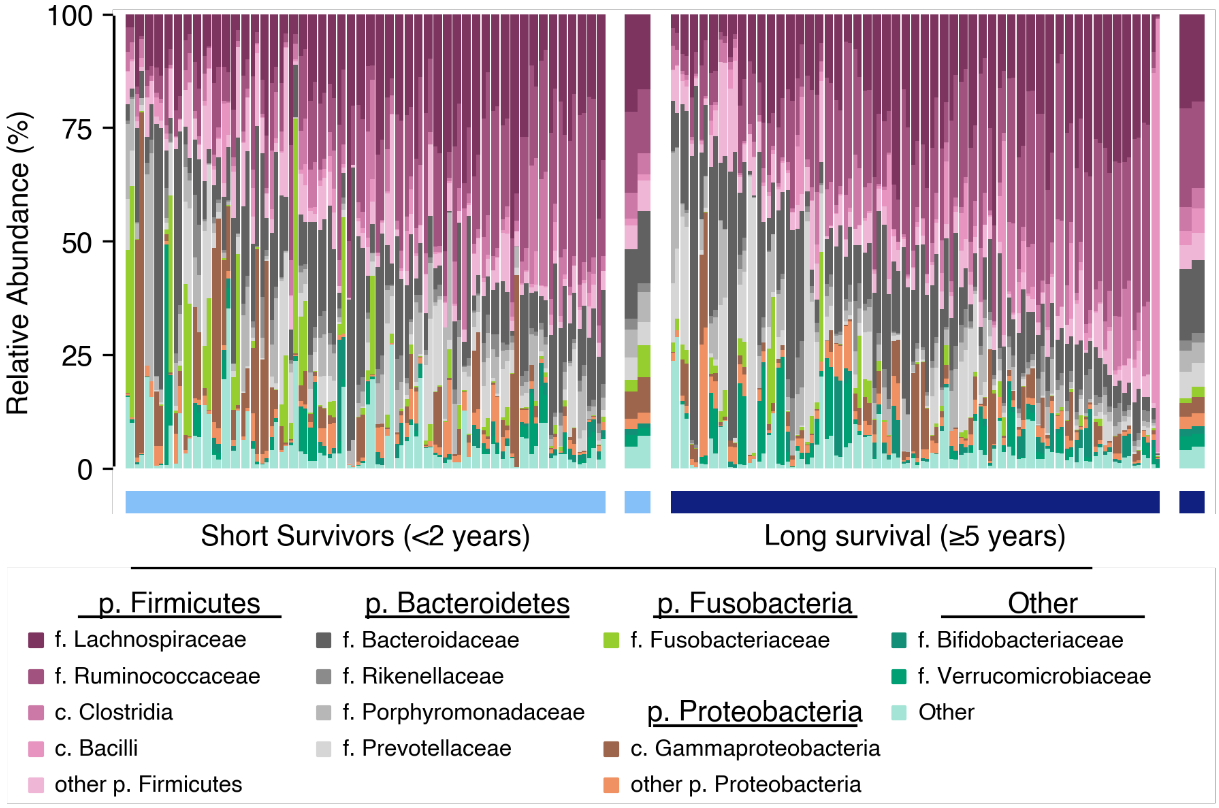


**Figure S1. A family level view of the microbiome**
Samples are sorted by individual donor with the normal tissue on the left and tumor on the right. Data is colored by taxonomic group (p: phylum level; c: class; f: family). The wide bars to the right of each distribution group show the average of the normal (left) and tumor (right) for all samples in the survival group.

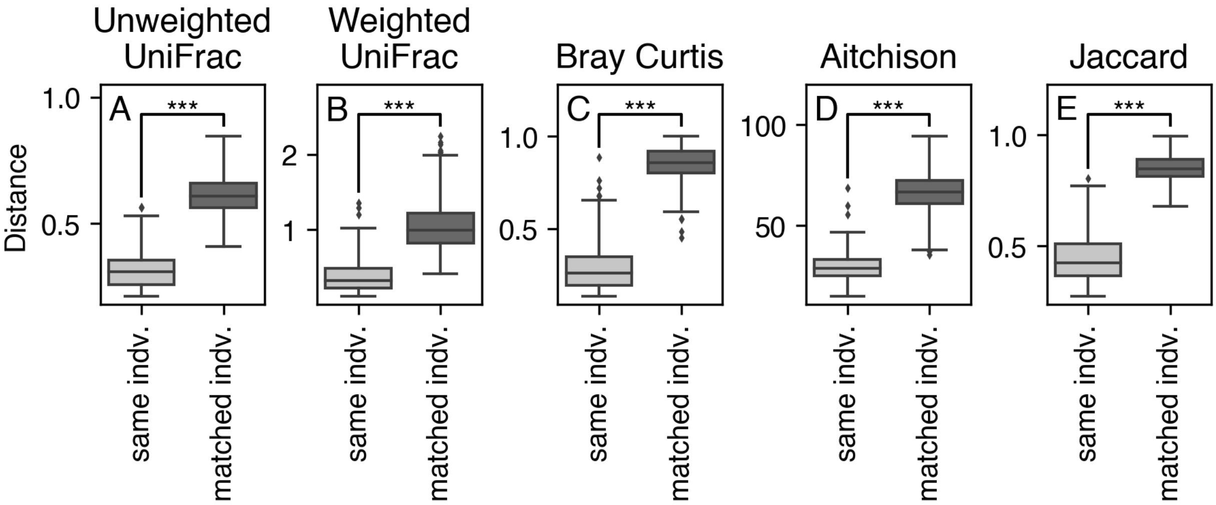


**Figure S2. There is a strong individual effect on the microbiome**
Beta diversity measured with (A) unweighted and (B) weighted UniFrac distances, (C) Bray Curtis dissimilarity, (D) Aitchison Distance, and (E) Binary Jaccard distance show within individual paired sample distances compared to the distance other samples matched on tissue type, anatomical location, surgery year, and survival. P-value from permutative t-test with 999 permutations; ***p=0.001, **p ≤ 0.01, *p ≤0.05.

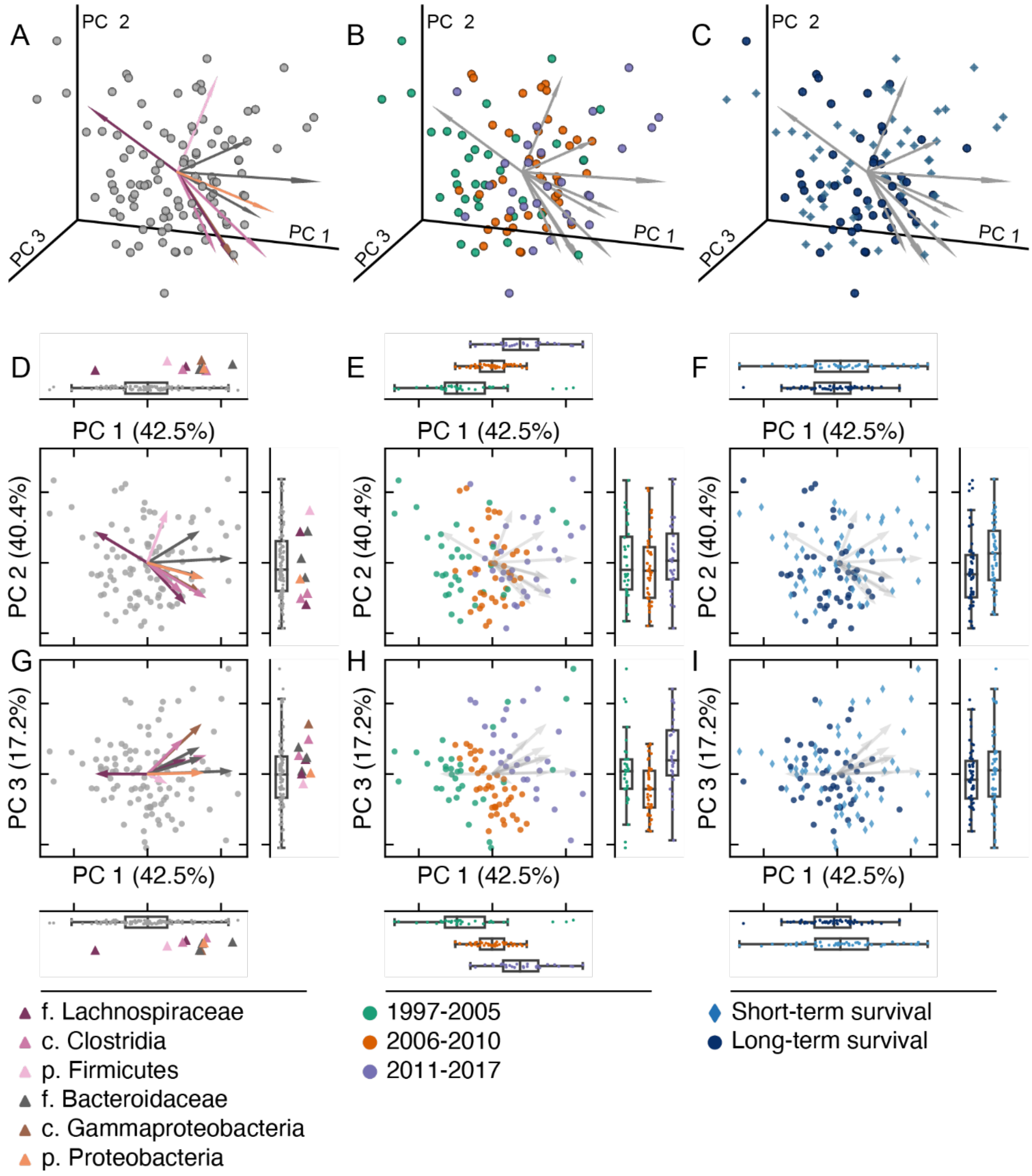


**Figure S3. Metadata predictors of the microbiome in Compositional Tensor Factorization.** Ordination plots show (A-C) three-dimension PCA projection plots, (D-F) PC 1 vs PC 2 and (G-I) PC 2 vs PC 3 for the per-subject ordination. Plots show (A,D,G) biplot features, (B,E,H) the period of surgery, and (C,F,I) the survival. Marginal axes show the distribution of samples along their respective axes.

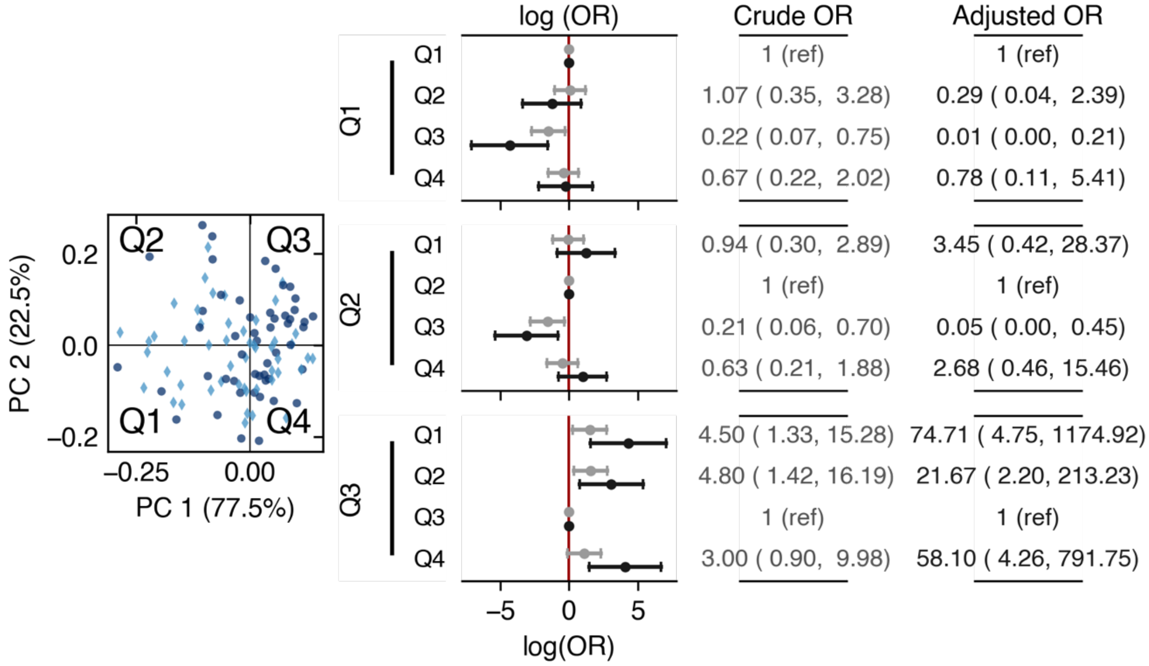


**Figure S4. Survival separates individuals by tumor-associated rPCA quadrant**
Left shows the quadrants in the rPCA space. Middle plot shows the log odds ratio (OR) for pairwise quadrants with 95% CI. Negative log OR indicates long survival, positive values indicate short survival. Red line indicates 0. Right shows the OR for the crude (light gray) and adjusted (dark gray) of short survival. The Adjusted OR is adjusted for age, sex, ASA score, tumor location, surgery year, TNM stage, and grade of differentiation.
